# Centhaquine Increases Stroke Volume and Cardiac Output in Patients with Hypovolemic Shock

**DOI:** 10.1101/2024.03.27.24304929

**Authors:** Aman Khanna, Krish Vaidya, Dharmesh Shah, Amaresh K. Ranjan, Anil Gulati

## Abstract

**Background:** Centhaquine is a resuscitative agent that acts on α2B adrenergic receptors to enhance venous return in hypovolemic shock. The effect of centhaquine on cardiac output in patients with hypovolemic shock has not been reported.

**Methods:** Trans-thoracic echocardiography was utilized to measure stroke volume (SV), cardiac output (CO), left ventricular outflow tract-velocity time integral (LVOT-VTI), left ventricular outflow tract diameter (LVOTd), heart rate (HR), left ventricular ejection fraction (LVEF), left ventricular fractional shortening (FS) and inferior vena cava (IVC) diameter before (0 min) and after centhaquine (0.01 mg/kg, iv infusion over 60 min) treatment (60 min, 120 min, and 300 min) in 12 randomly selected patients with hypovolemic shock enrolled in a prospective, multicenter, open-label phase IV clinical study (NCT05956418) of centhaquine in patients with hypovolemic shock.

**Results:** A significant increase in SV (mL) was observed after 60, 120, and 300 minutes of centhaquine treatment. CO (mL/min) increased significantly at 120 and 300 min despite a decrease in HR at these times. A significant increase in IVC diameter and LVOT-VTI (mL) at these time points was observed, which indicated increased venous return. The LVEF and FS did not change, while the mean arterial pressure (MAP, mmHg) increased in patients after 120 and 300 minutes of centhaquine treatment. Positive correlations between IVC diameter and SV (R^2^ = 0.9556) and between IVC diameter and MAP (R^2^ = 0.8928) were observed, which indicated the effect of centhaquine mediated increase in venous return on SV, CO, and MAP.

**Conclusions:** Centhaquine mediated increase in venous return appears to play a critical role in enhancing SV, CO, and MAP in patients with hypovolemic shock; these changes could be pivotal for reducing shock-mediated circulatory failure, promoting tissue perfusion, and improving patient outcomes.

**Trial registration:** The phase IV trial reported in this study has Clinical Trials Registry, India; ctri.icmr.org.in, CTRI/2021/01/030263; clinicaltrials.gov, NCT05956418.

## 1 Introduction

Hypovolemic shock decreases circulating blood volume and reduces stroke volume (SV), cardiac output (CO)[1], and tissue blood perfusion, leading to the possibility of organ failure and death. Managing hypovolemic shock is critical and involves prompt and targeted interventions to restore the circulating blood volume and increase organ perfusion using various fluids and vasopressors[1; 2]. Although the commonly used fluids (crystalloids or colloids) help compensate for the volume loss, patients often require vasopressors to achieve perfusion endpoints (e.g., CO and MAP)[3]. Vasopressors induce constriction of the blood vessels and aid the sympathetic system to increase blood pressure and CO, in an attempt to increase tissue perfusion[4]. However, the use of vasopressors (e.g., epinephrine, norepinephrine, dopamine, vasopressin, and angiotensin) remains debatable due to associated risks, including cardiac arrhythmias, decreased tissue perfusion, fluid extravasation, and organ failure [5].

At the normal physiological state, a significant amount of blood is pooled on the venous side (having high vascular capacitance) of the circulation; however, in hypovolemic shock, blood accumulation in the veins is further increased, and that a significant amount of blood does not participate in tissue perfusion[6]. It is of interest to divert the pooled venous blood towards the arterial side so that the circulating blood volume can be increased, which will help increase stroke volume (SV), cardiac output (CO), and mean arterial pressure (MAP), as well as increase tissue perfusion.

Centhaquine (2-[2-[4-(3-Methylphenyl)-1-piperazinyl]ethyl] quinoline citrate) is being developed to treat hypovolemic shock. It showed good tolerability and safety in healthy subjects with minimal adverse effects observed at almost ten times the therapeutic dose (NCT02408731). Clinical studies in hypovolemic shock patients (NCT04056065 and NCT04045327) found that centhaquine significantly increased HR and MAP, reduced lactate levels, and increased survival[7; 8]. The proposed mechanism of action of centhaquine is by acting on venous α2B-adrenergic receptors; it augments the blood return to the heart and increases SV[9]. In swine and rat models of hypovolemic shock, centhaquine significantly increased the SV, leading to increased CO and MAP [10; 11] and improved survival. The current study investigates centhaquine’s role in increasing cardiovascular variables (SV, CO, and MAP) by enhancing venous blood return in human patients with hypovolemic shock.

## 2 Materials and methods

### 2.1 Study Design

This pilot study was conducted on 12 randomly selected patients enrolled in a prospective, multicenter, open-label phase IV clinical study (NCT05956418) of centhaquine in patients with hypovolemic shock. At the baseline, demographic data, chest X-ray, electrocardiogram (ECG), and vital signs were recorded along with blood counts and chemistry. Patients received 0.01 mg/kg centhaquine by intravenous infusion over 60 min in 100 mL normal saline. Systolic and diastolic blood pressure (SBP and DBP) were recorded using a sphygmomanometer at baseline, hourly for the initial 48 hrs, and in between if needed. The mean arterial pressure [DBP + 1/3 (SBP – DBP)] and pulse pressure (SBP-DBP) were calculated.

The ongoing phase IV study is an open label study aimed at assessing the post-marketing efficacy of centhaquine. All patients in this study received centhaquine along with standard of care for hypovolemic shock. Data from the baseline value in this study was compared. Historical data on fluid resuscitation was also used to understand the superiority of resuscitation with centhaquine. The study duration for an individual patient was seven days or earlier (patient discharge).

#### 2.1.2 Echocardiographic measurements

Transthoracic echocardiography was utilized to assess SV, CO, HR, left ventricular ejection fraction (LVEF), and left ventricular fractional shortening (LVFS) before (0 min) and after centhaquine treatment (60 min, 120 min and 300 min). An expert technician conducted the echocardiography, and the detailed methodology is as follows -

2.1.2.1 LVOT diameter: A parasternal long-axis view was obtained to visualize the left ventricular outflow tract (LVOT) and the aortic valve. The view of the aortic valve opening and closing was ensured. The best view of the aortic valve at mid-systole (when the valves are wide open) was ensured, images were captured, and the distance near the aortic annulus at the base of the leaflets was measured using the tool. This measured distance was the diameter of the LVOT.

2.1.2.2 LVOT VTI (LVOT velocity time integral): The pulse-wave Doppler gate was aligned parallel to the LVOT in the Apical 5 Chamber view to obtain the best VTI tracing. Once the Doppler gate was positioned well, the pulse wave Doppler was activated. The automated LVOT VTI was calculated after tracing the outline of one of the systolic waveforms. The values for the LVOT VTI are denoted as the distance in centimeters (cm) and represent the distance that blood travels in one heartbeat.

2.1.2.3 Heart rate (HR): The specific points on the screen corresponding to individual heartbeats were marked, and the heart rate was automatically calculated by the echocardiography machine based on this input.

2.1.2.4 SV and CO: SV was calculated as the product of the LVOT diameter and the LVOT VTI. CO was calculated as the product of SV and HR.

2.1.2.5 LVEF and LVFS: LVEF was calculated via visual estimation by reviewing different echocardiography windows, as was done in an emergency setting. LVFS was calculated using the formula (%LVFS = [(LVDD-LVDS)/LVDD] × 100, where LVDD is the left ventricle diameter at diastole, and LVDS is the left ventricle diameter at systole, measured through echocardiography.

### 2.2 Patient population, consent, and regulatory oversight

The phase IV study’s detailed inclusion/exclusion criteria (NCT05956418) are provided at https://clinicaltrials.gov/study/NCT05956418. Adult hypovolemic shock patients aged ≥ 18 years with an SBP ≤ 90 mmHg upon presentation to the emergency room or ICU, receiving standard shock treatment, and having a blood lactate level >2.0 mmol/L were included in the study.

Informed consent from all enrolled patients or their legally authorized representative (LAR) was obtained verbally and in writing after communicating the study details.

The study adhered to the Harmonization of Technical Requirements for Registration of Pharmaceuticals for Human Use Guideline for Good Clinical Practice (ICH-GCP), the Helsinki Declaration, and local regulatory requirements.

### 2.3 Safety evaluation

Safety was evaluated by the study investigators based on adverse events (AEs), physical examination results, vital signs (including HR, SBP, DBP, body temperature, and respiratory rate), ECG, and clinical variables. Any AEs that occurred or worsened during or after centhaquine treatment were systematically recorded and coded by system organ class and preferred term using the latest version of the International Conference on Harmonization Medical Dictionary for Regulatory Activities.

### 2.4 Multivariate Imputation by Chained Equations (MICE)

Data not available (6.4%) were assessed as “missing values,” and they were imputed using “MICE,” which is a package that implements a method to address missing data by creating multiple imputations (replacement values) for multivariate missing data[12]. In this study, five patients had multiple imputations for missing echocardiography data. A scalar of 20, given the number of iterations and predictive mean matching “pmm,” was used.

### 2.5 Statistical analysis

The results are presented as the mean ± standard error of the mean (SEM). The statistical analysis was performed using GraphPad Prism 10.1.2 (GraphPad, San Diego, CA, USA). Parametric analysis was carried out using a one-way analysis of variance without assuming equal variances with a normal probability distribution. The post hoc Tukey’s multiple comparisons test was performed to estimate the significance of differences. p values < 0.05 were considered to indicate statistical significance at the 95% confidence level.

## 3 Results

### 3.1 Patient demographics, baseline characteristics, and volume of fluid administration during the first 5 hours of resuscitation

The demographics and baseline characteristics are shown in Table 1.

**Table 1.** Baseline characteristics of patients.

|  |  |
| --- | --- |
| Age (years) | 37.42 ± 3.68 |
| Body weight (kg) | 58.83 ± 1.57 |
| Height (cm) | 157.67± 1.2 |
| Body mass index (kg/m<sup>2</sup>) | 23.64 ± 0.22 |
| **Sex** |  |
| Men | 9 (75%) |
| Women | 3 (25%) |
| **Medical history** |  |
| Hypertension | 0 |
| Diabetes | 0 |
| Renal disorders | 0 |
| Respiratory disease | 1 (8.33%) |
| Ischemic heart disease | 0 |
| Liver fibrosis | 0 |
| Hepatitis (Altered SGPT) | 6 (50%) |
| Preeclampsia | 0 |
| **Reason for hypovolemic shock** |  |
| Gastroenteritis (vomiting, abdominal pain and or diarrhea) | 8(66.67%) |
| Enteric fever | 4(33.33%) |
| Dengue fever | 2 (16.67%) |
| Falciparum malaria | 1 (8.33%) |
| Acute appendicitis | 1 (8.33%) |
| **Clinical factors** |  |
| Systolic blood pressure (mmHg) | 73.87 ± 2.9 |

**Table 1. Baseline characteristics of patients**
|  |  |
| --- | --- |
| Diastolic blood pressure (mmHg) | 45.33 ± 1.64 |
| Heart rate (beats/min) | 108 ± 4.4 |
| ECG | Normal |
| Random blood glucose (mg/dL) | 104.24 ± 4.75 |
| Shock index | 1.51 ± 0.12 |
| MODS | 2.5 ± 0.38 |
| ARDS | 0.08 ± 0.083 |
| Respiratory rate (breaths/min) | 23.5 ± 0.23 |
| Body temperature (°F) | 102.3 ± 0.36 |
| Blood lactate (mmol/L) | 2.5 ± 0.06 |
| Base deficit (mmol/L) | -1.49 ± 0.06 |
| Hemoglobin (g/dL) | 12.16 ± 0.45 |
| Hematocrit (%) | 37.24 ± 1.26 |
| Creatinine (mg/dL) | 1.13 ± 0.15 |
| Glomerular filtration rate (mL/min/1.73 m<sup>2</sup>) | 83.43 ± 7.84 |
| pH | 7.21 ± 0.01 |
| pCO<sub>2</sub> (mmHg) | 33.17 ± 0.90 |
| PO<sub>2</sub>/FiO<sub>2</sub> | 376.75 ± 6.32 |

The data are presented as the mean ± SEM.

### 3.2 Centhaquine Increases Stroke Volume, Cardiac Output, and Mean Arterial Pressure

At baseline (0 min), the mean SV (mL) was 63.36 ± 4.06. At 60 min, 120 min, and 300 min, the mean SV (mL) was 78.07 ± 4.98 (Δ23.2%, p=0.0084), 83.51 ± 3.78 (Δ31.8%, p=0.0002), and 89.18 ± 3.71 (Δ40.74%, p<0.0001), respectively (Fig. 1A).

**Figure 1.**
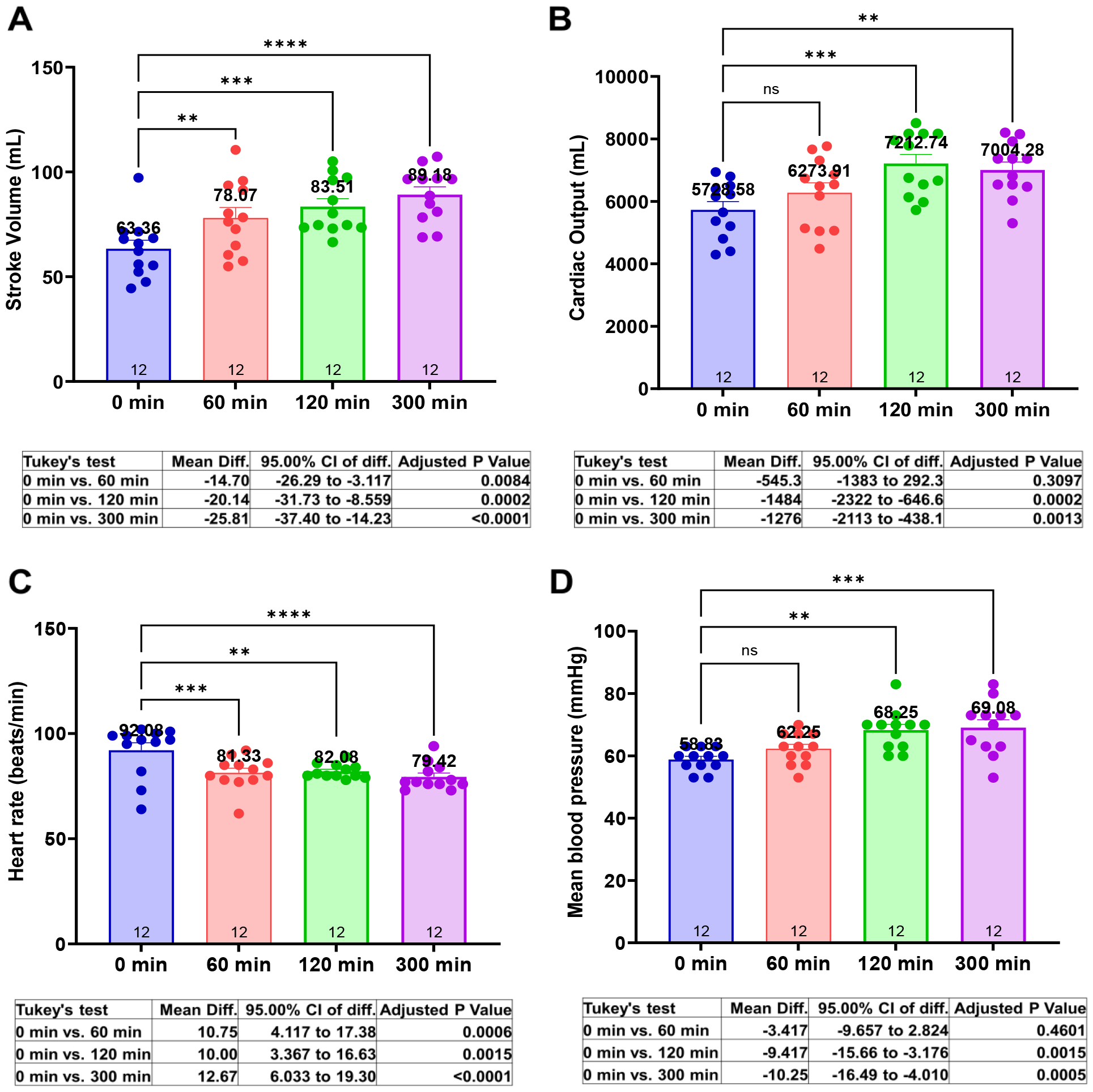
Effects of centhaquine on cardiovascular variables (SV, CO, HR, and MAP). * P<0.05, ** P<0.01, *** P<0.001, and **** P<0.0001 compared to 0 min. n= 12.

The mean CO (mL/min) at baseline was 5728.58 ± 263.4. At 60 min, 120 min, and 300 min, the mean SV (mL) was 6273.91 ± 318.33 (Δ9.52%, p=0.3097), 7212.74 ± 291.2 (Δ25.9%, p=0.0002), and 7004.28 ± 255.36 (Δ22.23%, p=0.0013), respectively (Fig. 1B).

At baseline, the mean HR (bpm) was 92.08 ± 3.55. At 60 min, 120 min, and 300 min, the mean HR (bpm) was 81.33 ± 2.23 (Δ11.69%, p=0.0006), 82.1 ± 0.96 (Δ10.97%, p=0.0015) and 79.42 ± 1.80 (Δ13.41%, p<0.0001), respectively (Fig. 1C).

MAP (mmHg) at baseline or 0 min was 58.89 ± 1.03. At 60 min, 120 min, and 300 min, the MAP values were 62.22 ±1.44 (Δ5.66%, p=.4601), 68.33 ± 1.86 (Δ16.04%, p=0.0015), and 69.27 ± 2.4 (Δ17.64%, p=0.0005), respectively (Fig. 1D).

### 3.3 Centhaquine Increases the Venous Return (increase in IVC Diameter)

The mean IVC diameter (cm) at baseline was 0.92 ± 0.04. A change in IVC diameter was observed after centhaquine treatment at 60, 120, and 300 min; the mean IVC diameter (cm) was 1.07 ± 0.03 (Δ15.94%, p=0.0091), 1.14 ± 0.02 (Δ25.00%, p<0.0001) and 1.14 ± 0.03 (Δ23.19%, p<0.0001), respectively (Fig. 2A).

**Figure 2.**
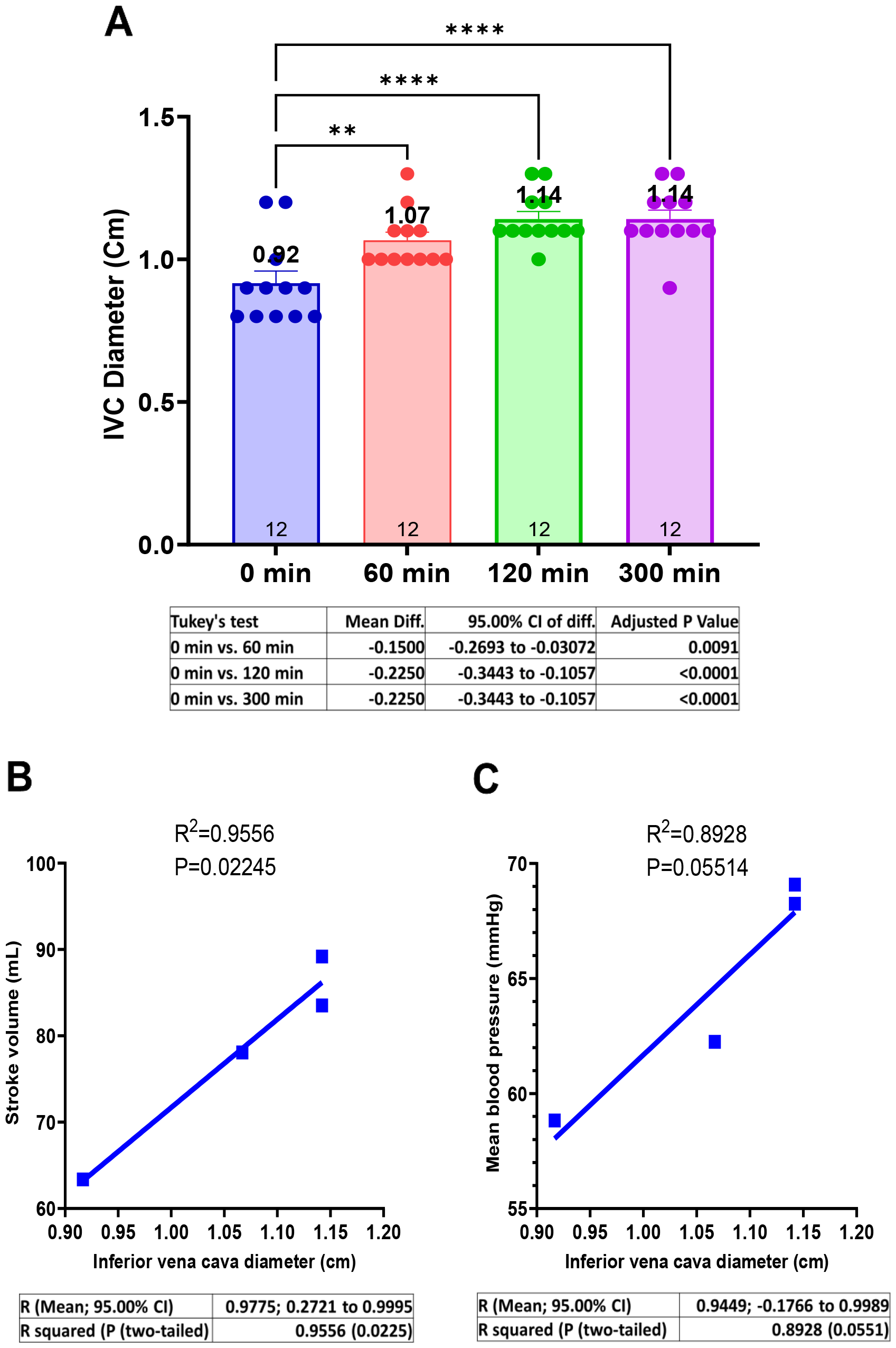
Effect of centhaquine on venous return (IVC diameter) and its correlation with SV and MAP. * P<0.05, ** P<0.01, *** P<0.001, and **** P<0.0001 compared to 0 min (A). n= 12.

The relationships between “IVC diameter and SV” and “IVC diameter and MAP” were evaluated. A direct correlation (R^2^ = 0.9556; p=0.02245) between IVC diameter and SV (Fig. 2B) and between IVC diameter and MAP was observed (R^2^ = 0.8928; p=0.05514) (Fig. 2C).

### 3.4 Centhaquine Increases LVOT-VTI (Blood Flow towards Aortic Annulus/Valve)

LVOT-VTI (cm) indicates the blood flow from the left ventricle of the heart towards the aorta. LVOT-VTI was 18.54 ± 1.11 at the baseline, while it was 21.97 ± 1.05 (Δ18.52%, p=0.0159) 24.14 ± 0.76 (Δ30.2%, p<0.0001), and 24.9 ± 0.8 (Δ34.15%, p<0.0001) at 60 min, 120 min, and 300 min, respectively (Fig. 3A). A direct correlation (R^2^ = 0.9796; p=0.01024) between LVOT-VTI and IVC diameter was observed (Fig. 3B). LVOT diameter and area remain unchanged (Fig. 3C and D) at these time points.

**Figure 3.**
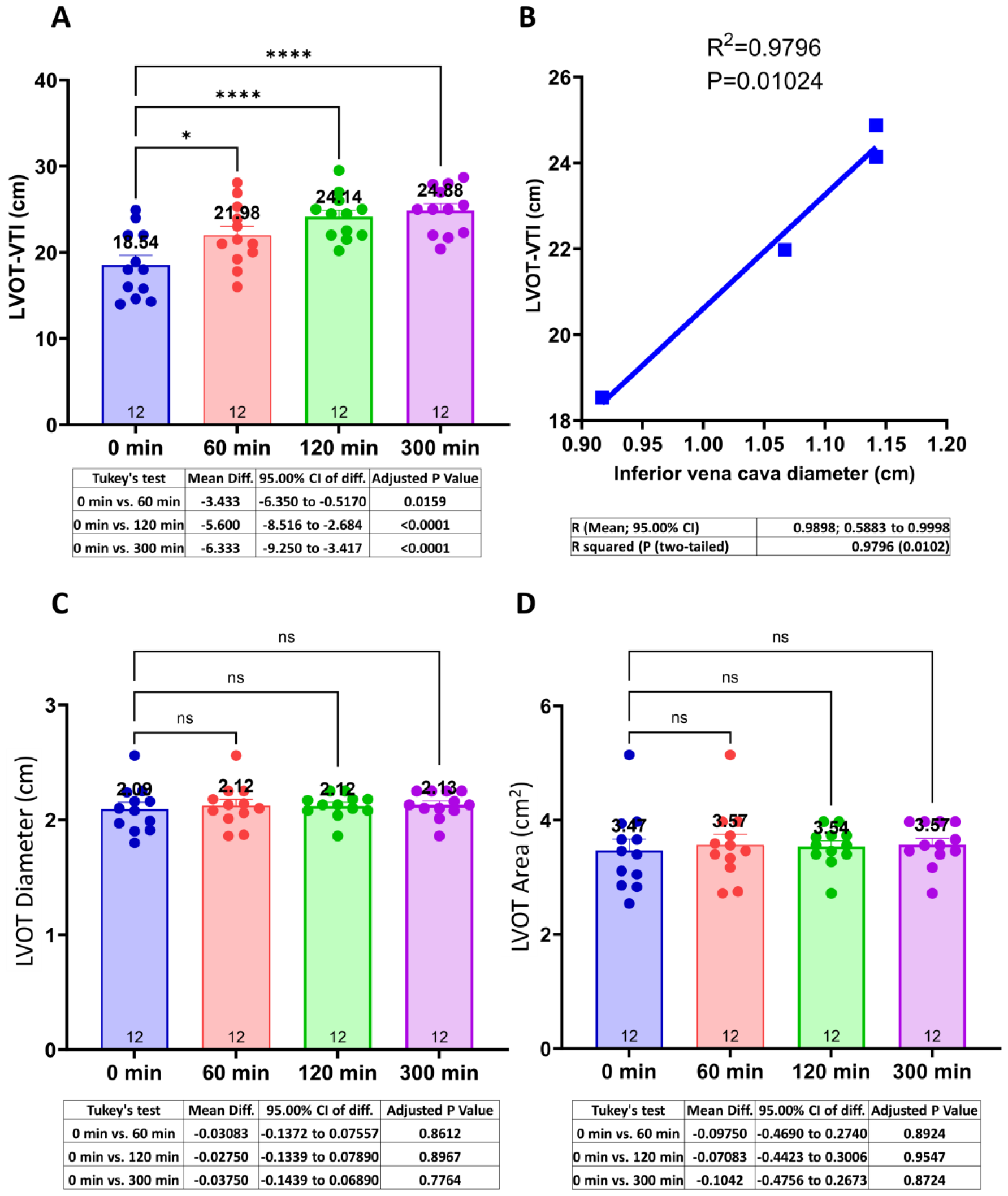
Effect of centhaquine on blood flow (LVOT-VTI) in LVOT to the aorta and its correlation with IVC diameter. * P<0.05, ** P<0.01, *** P<0.001, and **** P<0.0001 compared to 0 min (A). n= 12.

## 4 Discussions

The current study was conducted on 12 hypovolemic shock patients from a single cohort phase IV (NCT05956418) study; centhaquine increased SV and decreased HR (Fig. 1A and C) with ∼40% increase in SV, which is much higher compared to other studies relying on fluid infusion alone (∼10-25%) [13; 14]. Kumar et al. demonstrated a 15-25% increase in SV in healthy humans after infusion of 3 liters of normal saline at the rate of 1 liter per hour and observed cardiac inotropic effect of fluids with a ∼14% increase in LVEF. On the other hand, in our current study, only 746.12 ± 87.42 ml of fluid was required for 5 hours (∼150 ml per hour) of resuscitation (Table 2), and no change in LVEF and LVFS (suppl fig. 1A and B) was seen, indicating no effect on cardiac inotropy. Thus, centhaquine increases SV independent of the volume of fluids during resuscitation and does not affect cardiac inotropy. Hence, risks of fluid extravasation and cardiac arrhythmia are mitigated [15], which are associated with a higher volume of fluids and using vasopressors to treat shock[16; 17].

**Table 2.** Volume of fluids administered to patients before randomization and during the first 5 hours of resuscitation.

| Normal saline (mL) | Normal saline with dextrose (mL) | Total volume (mL) |
| --- | --- | --- |
| 345.7 ± 67.2 | 400.42 ± 32.42 | 746.12 ± 87.42 |

The data are presented as the mean ± SEM.

CO and MAP were increased (Fig. 1B, D, and Suppl Table 1) after centhaquine treatment despite reduced HR and unchanged total peripheral resistance (Suppl Fig. 2), indicating an impact of increased SV on arterial circulation and tissue perfusion. IVC diameter was also significantly increased (Fig. 2A), reflecting increased venous return and cardiac preload [18; 19; 20] after centhaquine treatment. A direct correlation between IVC diameter and SV (R^2^ = 0.9556) (Fig. 2B), and IVC diameter and MAP (R^2^ = 0.8928) (Fig. 2C) was observed, which indicated an effect of increased venous return on SV, CO, and MAP.

Furthermore, LVOT-VTI was significantly upregulated after centhaquine treatment, which indicated increased blood flow in the left ventricular outflow tract during systole, leading to enhanced SV[21; 22]. The increased blood flow towards LVOT could be attributed to increased chronotropy, cardiac inotropy, or venous return. After centhaquine treatment, however, patients were observed with decreased cardiac chronotropy and no change in inotropy (Suppl Fig. 1A and B); hence, increased venous return would be the main reason for the increased LVOT-VTI. The observed direct correlation between IVC diameter and LVOT-VTI ((R^2^ = 0.9796; p=0.01024) (Fig. 3B) has further supported the role of centhaquine mediated enhanced venous return in improving blood flow towards LVOT and enhancing SV.

Increased venous return and flow in LVOT would increase the blood volume in the arterial system, leading to increased MAP. Besides blood volume, vascular resistance is also vital for regulating blood pressure. Nonetheless, centhaquine treatment demonstrated no significant change in total peripheral resistance (Suppl Fig. 2), further supporting the role of centhaquine mediated increased venous return on arterial blood volume, causing an increase in MAP in patients with hypovolemic shock.

All patients treated with centhaquine in the study showed improved HR, respiratory rate, and body temperature. A reduction in serum lactate, base deficit, and an increase in PO2/FiO2 was observed in hypovolemic shock patients treated with centhaquine (Table 3). Improved patient outcomes were observed with decreased MODS (0.17 ± 0.11 at the time of discharge vs 2.5 ± 0.38 at the baseline), and stabilized hematological, biochemical, and serum electrolyte levels (Suppl Table 1). All 12 centhaquine treated patients recovered and were discharged at 3.1 ± 0.074 days. Thus, the efficacy and safety of centhaquine in hypovolemic shock patients is promising.

**Table 3.** Patient vital signs were recorded from day 1 (baseline) through days 3/4.

| Vitals | Baseline | After administration of centhaquine |  |  |
| --- | --- | --- | --- | --- |
|  | Day 1 | Day 2 | Day 3 | Day 4 |
| SBP (mmHg) | 73.83 ± 2.88 | 102.17 ± 1.17 | 110.00 ± 1.74 | 120.00 ± 1.67 |
| DBP (mmHg) | 45.33 ± 1.64 | 63.17 ± 2.47 | 69.17 ± 1.49 | 77.78 ± 1.47 |
| Heart rate (beats/min) | 108 ± 4.38 | 95.5 ± 3.27 | 92.42 ± 2.30 | 85.56 ± 1.45 |
| Respiratory rate (breaths/min) | 23.5 ± 0.23 | 21.83 ± 0.65 | 21.25 ± 0.64 | 20.11 ± 0.59 |
| Body temperature (°F) | 102.28 ± 0.36 | 99.51 ± 0.64 | 98.21 ± 0.30 | 97.57 ± 0.20 |
| Lactate (mM/L) | 2.5 ± 0.06 | 2.02 ± 0.03 | 1.67 ± 0.04 | 1.17 ± 0.05 |
| Base deficit (mM/L) | -149 ± 0.06 | -0.52 ± 0.09 | 0.38 ± 0.18 | 0.43 ± 0.04 |
| PO2/FiO2 | 376.75 ± 6.32 | 405.5 ± 1.74 | 419.45 ± 1.98 | 427.51 ± 3.05 |

The data are presented as the mean ± SEM. *DBP* diastolic blood pressure, *SBP* systolic blood pressure.

These findings align with studies in animal models of hypovolemic shock[10; 11; 23; 24; 25], leading to increased venous return, CO, MAP, and tissue perfusion and underscore the significance of venous return in managing shock[26], with centhaquine targeting this through α2B-AR agonism. Thus, the current study and other studies[22; 23; 24] highlight the importance of venous return for treating shock. The venous return is modulated through the venous tone, which is primarily regulated by the sympatho-adrenergic system, and thus, adrenergic signaling appears to be an important target in treating various types of shock[27; 28; 29; 30]. Interestingly, ARs are distributed distinctly in the arterial and venous systems and play a key role in coordinating the arterial and venous circulation. Most arteries and large veins (e.g., vena cava) are mainly regulated by α1/α2-ARs, while peripheral veins are regulated by α2B-ARs[25; 31; 32; 33]. The high abundance of α2B-ARs in the peripheral veins highlights their involvement in the constriction of these peripheral veins. The peripheral veins have relatively higher capacitance than central veins (58.95% vs 11.05%) and hence are vital players for regulating venous return and cardiac preload, which proportionately affects SV. Therefore, findings of the study demonstrating increased SV with increased IVC diameter indicate that centhaquine would be acting on the α2B-ARs present in the peripheral veins and inducing venoconstriction, which would mobilize the unstressed blood present in these veins towards the vena cava and leading to increased blood volume in IVC causing increase in its diameter (Fig. 2A).

However, further studies are required to elucidate which specific venous systems are affected after centhaquine treatment. Studies have shown that cutaneous and splanchnic veins, which together constitute the major blood reservoir in the body, respond to various factors, e.g., temperature, stress, arterial blood parameters, and blood pressure[27], elucidating the effect of centhaquine on the individual venous system would help explore its potential further for treatment of different types of shock involving circulatory failure in different regions. Moreover, further randomized controlled trials with larger cohorts are necessary to fully understand centhaquine’s effects on venous systems and its potential in treating different types of shock associated with circulatory failure.

## 5 Conclusions

The increased venous return induced by centhaquine plays a pivotal role in elevating SV, CO, and MAP mediated through increased LVOT-VTI and IVC diameter in patients experiencing hypovolemic shock. An increase in SV, CO, and MAP occurs concurrently with a decrease in heart rate without influencing the inotropic function of the heart. This unique combination of outcomes suggests that centhaquine has remarkable potential to mitigate circulatory failure associated with hypovolemic shock, thereby promoting blood flow and tissue perfusion and improving overall patient outcomes.

Understanding centhaquine’s distinctive mechanism of action raises the possibility of its development as a novel resuscitative agent not only for hypovolemic shock but also for other shock types (e.g., septic shock and vasodilatory shock) that share pathophysiological characteristics involving circulatory failure and hypotension. While further research and clinical studies are needed to fully elucidate centhaquine’s effectiveness and safety profile across diverse shock conditions, its ability to enhance venous return and cardiac performance without undesirable effects on heart rate or inotropy could be pivotal for the development of a highly effective and safer resuscitative agent for the treatment of shock.

## Supporting information

https://acrobat.adobe.com/id/urn:aaid:sc:VA6C2:5d288681-e7e8-4d52-ba1a-7d6856fd3872

## Data Availability

The anonymized patient datasets generated and/or analyzed during the study are available from the corresponding author on a reasonable request from a bona fide researcher/research group.

## 7 List of abbreviations

AR: Adrenergic Receptors
SV: Stroke Volume
CO: Cardiac Output
LVEF: Left Ventricular Ejection Fraction
FS: Fractional Shortening (Left Ventricular)
IVC: Inferior Vena Cava
LVOT: Left Ventricular Outflow Tract
VTI: Velocity Time Integral
LVDD: Left Ventricle Diameter at Diastole
LVDS: Left Ventricle Diameter at Systole
DBP: Diastolic Blood Pressure
HR: Heart Rate
MAP: Mean Arterial Pressure
SV: Stroke Volume
SVR: Systemic Vascular Resistance
SBP: Systolic Blood Pressure
SOC: Standard of Care
MICE: Multivariate Imputation by Chained Equations
MODS: Multiple Organ Dysfunction Syndrome
ARDS: Acute Respiratory Distress Syndrome

## 8 Declaration

### Ethics Approval and Consent to Participate

The study was conducted in compliance with the Harmonisation of Technical Requirements for Registration of Pharmaceuticals for Human Use Guideline for Good Clinical Practice (ICH-GCP), the Helsinki Declaration, and local regulatory requirements. The study protocol (PMZ-2010/CT-4.1/2019), version 1.0/ dated October 16, 2019, was approved by the Drugs Controller General of India (DCGI), Directorate General of Health Services, Ministry of Health & Family Welfare, Government of India (DCGI CT NOC. No.: CT/ND/110/2020). Besides, each institutional ethics committee reviewed and approved the study protocol before initiating patient enrolment. The trial was registered at the Clinical Trials Registry, India (CTRI/2021/01/030263), and the United States National Library of Medicine, ClinicalTrials.gov (NCT05956418). Each site’s ethics committee was informed of any protocol deviation, amendment, subject exclusion or withdrawal, and serious adverse events (SAE). (Details of the participating site for this study are in Suppl Table 2)

### Consent

The patients included in this study were in a state of life-threatening shock; therefore, for patients who were not fit to give consent themselves at the time of treatment initiation, informed consent was obtained from their legally authorized representative (LAR). The investigator verbally, as well as in writing, informed the patient or LAR of the details of the study relevant to a decision about whether to participate in the study.

No patient identifiable data is present in this article.

## 10 Author Contributions

Conceptualization: A.G., and D.S.; Data curation: A.G., A.K.R. and D.S.; Formal analysis: A.K.R., A.G. and D.S.; Investigation: A.K., K.V.; Methodology: A.G., D.S. and A.K.; Project administration: A.K. and D.S.; Visualization: A.G. and A.K.R.; Writing–original draft: A.K.R.; Writing–review & editing: A.K.R., A.G. and D.S. All authors read and approved the final manuscript.

## 12 Acknowledgments

We acknowledge Sunil Gulati, Pharmazz Inc., USA, for imputing missing data using a package “Multivariate Imputation by Chained Equations (MICE),” which implements a method to address missing data by creating multiple imputations (replacement values) for multivariate missing data.

