## Supplementary material for "Centhaquine Increases Stroke Volume and Cardiac Output in Patients with Hypovolemic Shock": https://acrobat.adobe.com/id/urn:aaid:sc:VA6C2:5d288681-e7e8-4d52-ba1a-7d6856fd3872

**Supplementary Figure 1.** Effects of centhaquine on LVEF and LVFS. P > 0.05 = ns (not significant) compared to 0 min. n= 12.

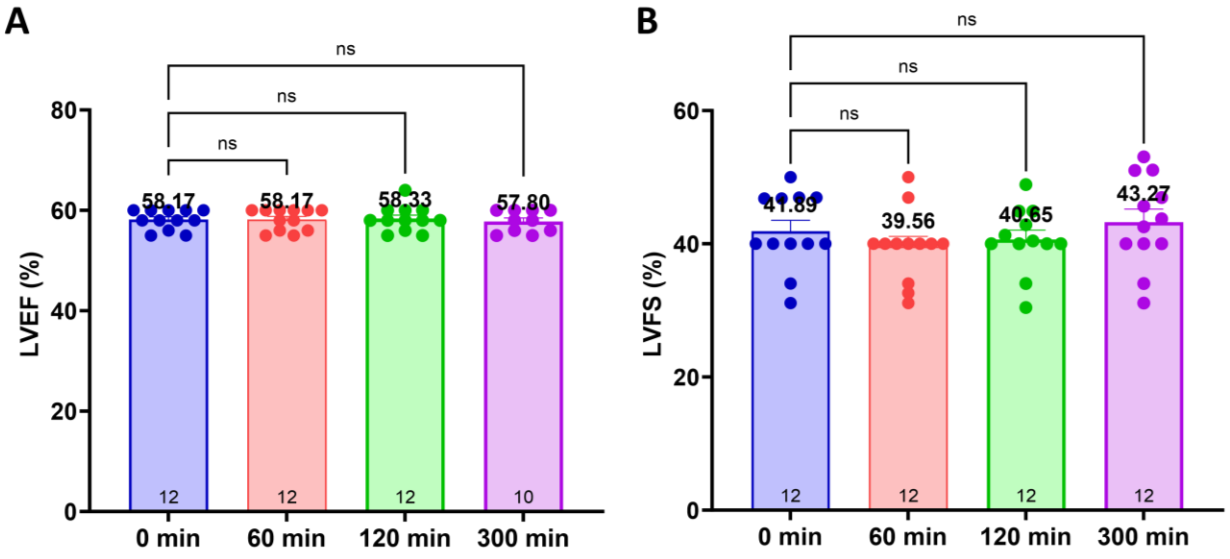


| Tukey's test | Mean Diff. | 95.00% CI of diff. | Adjusted P Value |
|---|---|---|---|
| 0 min vs. 60 min | 0.000 | -0.9711 to 0.9711 | >0.9999 |
| 0 min vs. 120 min | -0.1667 | -1.138 to 0.8044 | 0.9660 |
| 0 min vs. 300 min | 0.1444 | -0.8893 to 1.178 | 0.9811 |

| Tukey's test | Mean Diff. | 95.00% CI of diff. | Adjusted P Value |
|---|---|---|---|
| 0 min vs. 60 min | 2.329 | -3.191 to 7.849 | 0.6672 |
| 0 min vs. 120 min | 1.242 | -4.279 to 6.762 | 0.9287 |
| 0 min vs. 300 min | -1.379 | -6.899 to 4.142 | 0.9056 |

**Supplementary Figure 2.** Effects of centhaquine on vascular resistance (total vascular resistance). P > 0.05 = ns (not significant) compared to 0 min. n= 12.

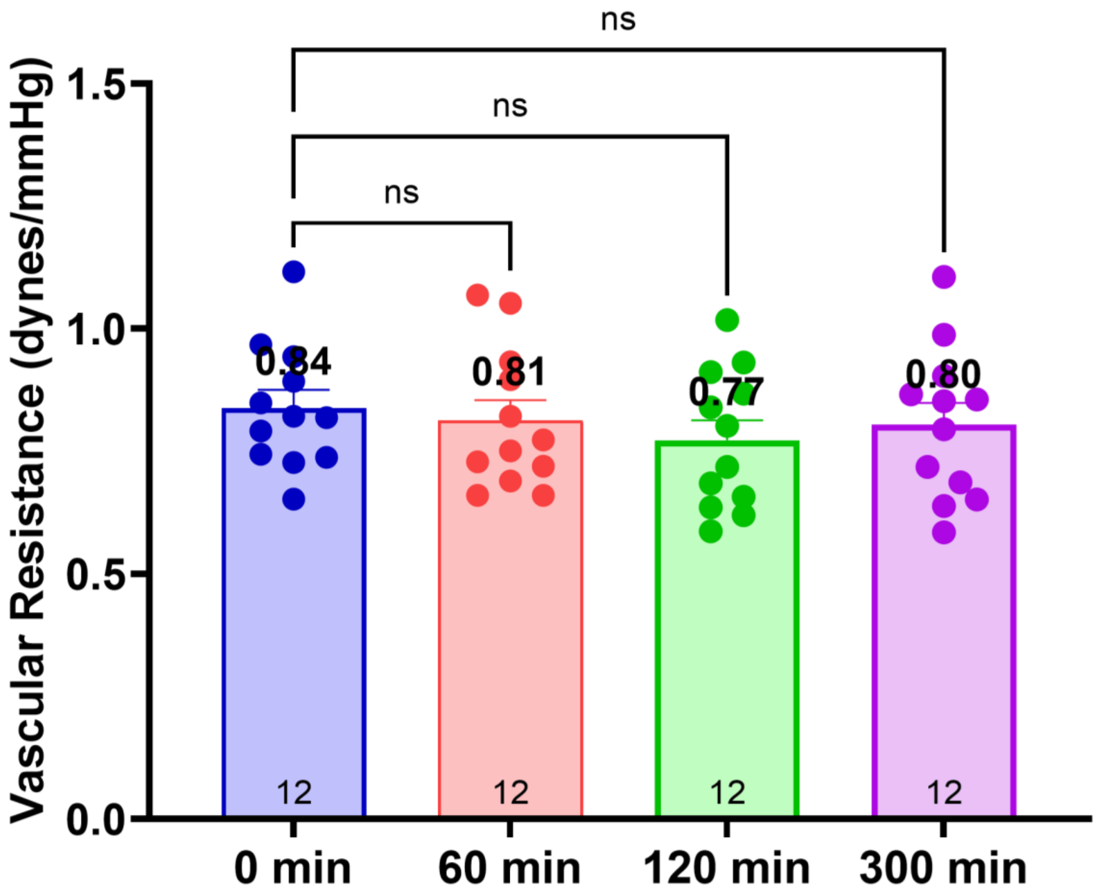


| Tukey's test | Mean Diff. | 95.00% CI of diff. | Adjusted P Value |
|---|---|---|---|
| 0 min vs. 60 min | 0.02557 | -0.09591 to 0.1470 | 0.9405 |
| 0 min vs. 120 min | 0.06575 | -0.05573 to 0.1872 | 0.4700 |
| 0 min vs. 300 min | 0.03438 | -0.08709 to 0.1559 | 0.8693 |

**Suppl Table 1. Hematological, biochemical, and serum electrolyte levels**

| | **Day 1 (baseline)** | **Day 3/4** |
|---|---|---|
| **Hematology** | | |
| Hemoglobin (g/dL) | 12.16 ± 0.45 | 12.11 ± 0.4 |
| Hematocrit (%) | 37.24 ± 1.26 | 36.7 ± 1.02 |
| Red blood cells ($10^6/mm^3$) | 4.60 ± 0.15 | 4.58 ± 0.13 |
| White blood cells ($/mm^3$) | 10692.50 ± 2226.16 | 9176.01 ± 3097.34 |
| Neutrophils (%) | 74.08 ± 3.356 | 66.50 ± 1.323 |
| Lymphocytes (%) | 19.17 ± 2.72 | 28.00 ± 1.32 |
| Monocytes (%) | 5.58 ± 1.11 | 3.67 ± 0.81 |
| Eosinophils (%) | 1.50 ± 0.15 | 1.83 ± 0.11 |
| Basophils (%) | 0.00 ± 0.00 | 0.00 ± 0.00 |
| Reticulocytes (%) | 0.97 ± 0.062 | 0.90 ± 0.038 |
| Mean corpuscular volume (fL) | 81.08 ± 1.687 | 80.33 ± 1.360 |
| Mean corpuscular hemoglobin (Pg) | 26.41 ± 0.512 | 26.44 ± 0.521 |
| Platelets ($/mm^3$) | 163333.33 ± 22663.658 | 194750.00 ± 16502.123 |
| **Lipid profile** | | |
| Triglyceride (mg/dL) | 115.58 ± 6.34 | 118.58 ± 6.12 |
| Total cholesterol (mg/dL) | 166.00 ± 6.314 | 165.13 ± 5.138 |
| High-density lipoprotein (mg/dL) | 46.78 ± 4.335 | 42.02 ± 0.512 |
| Low-density lipoprotein (mg/dL) | 100.28 ± 6.340 | 99.50 ± 5.176 |
| Very-low-density lipoprotein (mg/dL) | 23.12 ± 1.267 | 23.72 ± 1.218 |
| **Kidney function** | | |
| Serum creatinine (mg/dL) | 1.13 ± 0.148 | 1.01 ± 0.081 |
| Blood urea nitrogen (mg/dL) | 16.20 ± 2.115 | 14.90 ± 1.934 |
| Glomerular filtration rate (ml/min/1.73 $m^2$) | 83.43 ±7.84 | 95.01 ±7.81 |
| **Liver function** | | |
| Alanine aminotransferase (U/L) | 61.74 ± 8.710 | 47.03 ± 4.163 |
| Aspartate aminotransferase (U/L) | 57.43 ± 7.884 | 43.88 ± 3.989 |
| Serum bilirubin (mg/dL) | 0.99 ± 0.064 | 0.87 ± 0.023 |
| Alkaline phosphatase (IU/L) | 119.80 ± 7.691 | 110.68 ± 5.344 |
| Serum albumin (g/dL) | 3.71 ± 0.060 | 3.65 ± 0.031 |
| Blood glucose (mg/dL) | 104.24 ± 4.746 | 95.32 ± 8.360 |

| | Day 1 (baseline) | Day 3/4 |
|---|---|---|
| **Serum electrolyte** | | |
| Sodium (mmol/L) | 138.08 ± 2.113 | 140.48 ± 1.031 |
| Potassium (mmol/L) | 3.96 ± 0.096 | 4.10 ± 0.054 |
| Calcium (mmol/L) | 8.77 ± 0.107 | 8.82 ± 0.094 |
| **Arterial blood gases** | | |
| pH | 7.21 ± 0.01 | 7.36 ± 0.00 |
| $pCO_2$ (mmHg) | 33.17 ±0.90 | 37.92 ±1.00 |
| $paO_2$ /$FiO_2$ | 376.75 ± 6.321 | 427.78 ± 2.475 |

The data are presented as the mean ± SEM. *pH* power of hydrogen, $paO_2$ partial pressure of oxygen, $pCO_2$ partial pressure of carbon dioxide, and $FiO_2$ fraction of inspired oxygen.

**Suppl Table 2. Study Site Details**

| Site No. | Name of Study Site | Name of Ethics Committee | EC Submission Date | EC Approval date |
|---|---|---|---|---|
| 12 | Aman Hospital and Research Centre<br><br>15 Shashwat, Opp, E.S.I Hospital Sarabhai, Gotri Road, Vadodara, Gujarat-390021, India | Institutional Ethics Committee<br><br>Aman Hospital and Research Centre<br><br>15 Shashwat, Opp, E.S.I Hospital Sarabhai, Gotri Road, Vadodara, Gujarat-390021, India | 07 Nov 2022 | 30 Nov 2022 |
